# Multivariate Analysis Of Histopathological And Immunohistochemical Prognostic Factors In Endometrial Carcinoma. A Retrospective Pilot Study Of An Italian Regional Referral Center

**DOI:** 10.1101/2020.12.09.20245779

**Authors:** Michele Paudice, Giulia Scaglione, Chiara Maria Biatta, Fabio Barra, Marianna Riva, Bruno Spina, Gabriele Gaggero, Ezio Fulcheri, Simone Ferrero, Valerio Gaetano Vellone, Gyn DMT

**Affiliations:** Dept. of Integrated Diagnostic and Surgical Sciences (DISC), University of Genoa, Italy; Dept. of Neurosciences, Rehabilitation, Ophthalmology, Genetics, Maternal and Child Health (DINOGMI), University of Genoa, Italy; Pathology Unit, San Martino Hospital, Genoa, Italy; Fetal and Perinatal Pathology Unit, Istituto Giannina Gaslini, Genoa; Obstetrics & Gynecology Clinic, San Martino Hospital, Genoa, Italy; Pathology University Unit, San Martino Hospital, Genoa, Italy

**Author notes:** Corresponding Author: Valerio Gaetano Vellone MD, PhD, Associate Professor of Pathology-Consultant Gynecologic and Fetal Pathologist-Dept. Of Integrated Surgical and Diagnostic Sciences (DISC). University of Genoa, Via A. De Toni 14 16132 Genoa, Italy. these authors equally contributed in the redaction of this paper (Michele Paudice, Giulia Scaglione). GYN DMT, a multidisciplinary team of specialist physicians of Ospedale Policlinico San Martino, Genoa, involved in diagnosis and treatment of gynecologic malignancies composed by Serafina Mammoliti (Oncology), Cinzia Caroti (Oncology Clinic), Sergio Costantini (Obstetrics & Gynecology Clinic), Mario Valenzano Menada (Obstetrics & Gynecology Clinic), Melita Moioli (Obstetrics & Gynecology Clinic), Maria Grazia Centurioni (Obstetrics & Gynecology), Franco Alessandri (Obstetrics & Gynecology), Luca Dogliotti (Radiology), Maurizio Cosso (Radiology), Michela Marcenaro (Radiation Oncology), Flavio Giannelli (Radiation Oncology), Stefano Di Domenico (General Surgery) and Franco De Cian (General Surgery).

**Keywords:** endometrial carcinoma, prognosis, morphology, immunohistochemistry

## Abstract

**Background:** to investigate endometrial carcinoma prognostic value of some histopathological and immunohistochemical factors, fairly easily accessible in every routinely pathology lab set.

**Methods:** we considered patients affected by endometrial carcinoma with available clinical and radiological follow-up data after radical hysterectomy (S. Martino Polyclinic Hospital, Genoa, Italy, period 1/1/2013 - 1/7/2016). We analyzed the following histopathological items: histotype, stage (FIGO), type of infiltration (infiltrative/espansive), desmoplasia, intratumoral necrosis, tumor infiltrating lymphocytes and lymph vascular spaces invasion. Moreover, each case has been investigated with a panel of immunohistochemistry including estrogen receptor α, progesteron receptor, Ki67, p53, β-catenin, e-cadherin, bcl-2 and cyclin D1. Primary endpoints were disease free survival and overall survival.

**Results:** out of 99 cases eligible for our purpose, we found 69 low-grade endometrioid, 8 high-grade endometrioid and 22 other high-grade endometrial carcinomas. Disease free survival multivariate analysis showed a strong significant correlation between poor prognosis and advanced stage (p=0.0042). Advanced stage (p=0.0003) and presence of desmoplasia (p=0.04) resulted significantly correlated to a worse prognosis in overall survival multivariate analysis. In univariate model, the non-endometrioid histotype was significantly correlated with an unfavorable prognosis when compared to the endometriod type. Same for progesteron receptor low expression.

**Conclusion:** the multivariate analysis confirmed the central prognostic role of stage in endometrial carcinoma. Moreover, other immunohistochemical markers in univariate analysis, have confirmed their easily reproducible usefulness, well integrating the recent TGCA molecular classification.

## Introduction

Endometrial carcinoma (EC) is the fifth most common cancer in women. The incidence is growing, simultaneously with the increase of specific tumor-related risk factors, such as advanced age, nulliparity, obesity and tamoxifen therapy. Recent evidences show also an increased prevalence of aggressive subtypes and advanced-stage disease.^1,2^

EC has been categorized into two major classes (Bokhman 1983), based on clinical– pathological correlations. EC type I, namely endometrioid EC (EEC), represents the majority of sporadic endometrial carcinomas (70-80%), estrogen-related and generally with a good clinical outcome. EC type II, or non-endometrioid EC (NEEC), is less frequent (about 10–20% of EC), unrelated to estrogen exposure, but more aggressive.^3^

In 2013, TCGA (Cancer Genome Atlas Research Network) reclassified EC in 4 novel molecular classes with a significant correlation with prognosis.^4^ Each group of EC is characterized by different development and progression based on distinct molecular mechanisms of oncogenesis, reflecting the presence of type-specific molecular alterations. However histopathological and immunohistochemical factors still have a crucial role in patient-related risk stratification, given that the latter classification is still far from a routinely lab use. Indeed, different studies in literature are trying to integrate the molecular classification with immunohistochemical (IHC) markers.^5,6^

Thereupon, we evaluated the prognostic relevance of morphological and IHC parameters already well investigated for EC, being some of them suggested to be present in pathological report.^7^ Regarding IHC markers we considered a panel of different antibodies, all individually studied in literature, and we evaluated their prognostic role when combined together.

The prognostic value of estrogen (ER) and progesterone receptors (PR) is well established in literature.^8,9,10,11^ Some studies associate ER and PR double loss to poor prognosis, even in low grade subtype.^8,12^ Hormonal therapy is applied as conservative treatment in a fertility-sparing setting or as palliative in old or advanced stage patients.^13^ However, no recommendation about ER and PR IHC analysis is present in any clinical guideline.^14^

TP53 oncosuppressor gene (chromosome 17) encodes a transcriptional factor involved in cell cycle arrest and apoptosis. TP53 mutation is a substantial prognostic biomarker, predicting unfavorable outcome, usually related to serous carcinoma.^15^ P53 sequencing is difficult to apply in routinary laboratory setting, thus the relatively less expensive p53 IHC stain is used as surrogate of mutation analysis.^16^

Ki67 is a nuclear antigen expressed by proliferating cells (phases G1, S, G2, mitosis), but absent in resting cells (G0). High Ki67 expression is related to a more aggressive behavior of cancer.^17,18^

One EEC subgroup is driven by CTNNB1 mutation with an unfavorable prognostic value and morphologically associated to low grade and squamous differentiation. CTNNB1 (exon 3) encodes a cell-cell adhesion protein β-catenin. In IHC, its mutation corresponds to a nuclear staining, whenever β-catenin expresses a cell membrane staining in normal endometrium. Although usually utilized as IHC surrogate for exon 3 mutation, the β-catenin nuclear accumulation has an insufficient concordance with CTNNB1 sequencing.^19,20^ Likewise, CDH1 (exon 16) oncosuppressor gene, encoding for e-cadherin, is related to cell cohesiveness. Low e-cadherin is related to tumor cells exfoliation and high risk of peritoneal metastasis. 60% of EC type 2 and 22% of EC type 1 harbor this mutation.^21^

BCL2 (exon 14) is a proto-oncogene which encodes bcl-2, a protein with anti-apoptotic activity. Loss of BCL2 is associated with independent negative prognostic factors, such as a greater myometrial invasion, aggressive histotype, loss of expression of PR and advanced FIGO stage. Studies showed a correlation between loss of bcl-2 and risk of lymph node metastasis and recurrence.^22,23,24^ Others have shown that the genes regulating apoptosis (BCL2, BAX, caspase 3) seem apparently involved in the shift from simple to complex hyperplasia and to adenocarcinoma.^25^

CCND1 (chromosome 11) is a proto-oncogene which encodes cyclin D1, and it is more typical of EC type 2.^21^ In EC, cyclin D1 overexpression has a negative prognostic value, and it is related to lymph node involvement. Rarely β-catenin and cyclin D1 are overexpressed together. Some studies showed that cyclin D1 alteration could be an early event in endometrial carcinogenesis.^26,27^

## Materials and methods

We considered patients treated with radical hysterectomy for EC in our institution from 1/1/2013 to 1/7/2016. Only cases without neoadjuvant chemotherapy or hormonal therapy were selected. Moreover, patients without a complete follow-up were excluded. Patients underwent a previous diagnostic hysteroscopy and intraoperative frozen section analysis at the time of surgery. Bilateral pelvic lymph node dissection was performed in case of intraoperative diagnosis of high grade EC or low-grade EC infiltrating more than 50% of the myometrial wall.

All the surgical specimens have been routinely fixed and processed to obtain 3 µm-thick histological sections, finally stained with hematoxylin/eosin (HE). All the cases have been investigated with a panel of IHC stain including ERα, PR, ki67, p53, β-catenin, e-cadherin, bcl-2, cyclin D1. The histopathological examinations have been reported using an institutional protocol.

For IHC we used an automatic immunostainer Benchmark XT (Ventana Medical Systems SA, Strasbourg, France). Antigen-retrieval was obtained with citrate buffer (pH 6) at 90°C for 30 minutes, incubated in primary antibody for 1 hour at 37°C followed by the addition of the polymeric detection system Ventana Medical System Ultraview Universal DAB Detection Kit, counterstained with modified Gill’s hematoxylin and mounted in Eukitt.

The tested antibodies are described in table 1.

**Table 1.** List of antibodies with specific clone

| **Marker** | **clone** | **manufacturer** | **dilution** |
| --- | --- | --- | --- |
| **ER** | 6F11 | Ventana | pre-diluted |
| **PR** | 1E2 | Ventana | pre-diluted |
| **Ki-67** | 30-9 | Ventana | pre-diluted |
| **p53** | DO-7 | Ventana | pre-diluted |
| **ciclin D1** | SP4-R | Ventana | pre-diluted |
| **bcl-2** | 124 | Ventana | pre-diluted |
| **β-catenin** | 14 | Ventana | pre-diluted |
| **e-cadherin** | 36 | Ventana | pre-diluted |

In brief: for all the proposed molecular markers, the staining index (SI), accounting the percentage (%) of positive tumor cells, have been evaluated by two pathologists working separately and in blind. At least 500 tumor cells have been evaluated, any intensity of staining from weak (1+) to intense (3+) has been considered as positive. Any discrepancy has been discussed at a multiheaded microscope to a final decision.

On the base of the distribution of the SI, we dichotomized the study population as follow: ER≤20% was considered as ER low, while ER>20% as ER high. The same classification has been proposed for PR and cyclin D1. On the base of the mean value of the study population patients with Ki67<45% have been classified as Ki67 low while patient with Ki67≥45% have been classified as Ki67 high. Cases with p53≥10% have been considered p53 positive. For e-cadherin and β-catenin values ≤80% were considered low. (Additional Material 1)

The clinical, pathological and IHC data of the patients enrolled in the study were entered in a Microsoft Excel © spreadsheet.

Discrete variables were compared using the χ^2^ test; continuous variables were compared using Kruskall-Wallis test. Correlation between continuous variables was studied with the Spearman Rank Correlation Coefficient. Survival univariate analysis was studied with Kaplan-Meier survival curves, survival and hazard multivariate analysis was studies with Cox-Model; the significance was confirmed with the Log Rank Test. For statistical computation MedCalc© and OriginPro® programs were used. In all cases a degree of significance of 95% was chosen. p<0.1 was considered of borderline significance. In the tables continuous numeric variables are expressed as mean ± standard deviation while categorical variables as the number of observed cases (percentage). The parameters we chose for multivariate analysis were those with p<0.1 in univariate analysis.

## Results

### 1-General features of the study population

Of the total 151 cases, 99 (65.56%) were eligible according to selection criteria.

The clinical, histopathological and immunohistochemical features of the study population are summarized in Table 2.

**Table 2.** Clinical, histopathological and IHC features of the study population

| Categorical variables |  |  |  |  |  | Continuous variables |  |  |
| --- | --- | --- | --- | --- | --- | --- | --- | --- |
| Histology | N | % | Stage (FIGO) | N | % |  | Mean | SD |
| **Type 1 (EEC)** | 77 | 77.78 | **LOCAL** | 73 | 73.74 | **Age** | 68.49 | 11.26 |
| Low-grade | 69 | 69.70 | IA | 53 | 53.54 | **ER (%)** | 65.98 | 33.94 |
| High-grade | 8 | 8.08 | IB | 15 | 15.15 | **PR (%)** | 53.51 | 36.11 |
| **Type 2 (NEEC)** | 22 | 22.22 | II | 5 | 5.05 | **Ki67 (%)** | 45.05 | 22.05 |
| Carcinosarcoma | 4 | 4.04 | **ADVANCED** | 26 | 26.26 | **p53 (%)** | 22.26 | 35.30 |
| Clear cell | 2 | 2.02 | IIIA | 7 | 7.07 | **e-cadherin (%)** | 96.60 | 11.96 |
| Mixed | 8 | 8.08 | IIIB | 3 | 3.03 | **β-cate nin (%)** | 95.10 | 10.90 |
| Serous | 6 | 6.06 | IIIC1 | 11 | 11.11 | **bcl-2 (%)** | 26.09 | 28.95 |
| Undifferentiated | 2 | 2.02 | IIIC2 | 5 | 5.05 | **cyclin D1 (%)** | 26.75 | 28.37 |
| **Follow-up Disease Free Survival (DFS)** | N | % | **Follow-up Overall Survival (OS)** | N | % | **Follow-up OS (months)** | 25.98 | 12.10 |
| No Relapse | 84 | 84.85 | Alive | 89 | 89.90 | **Follow-up DFS (months)** | 24.95 | 12.35 |
| Relapse | 15 | 15.15 | Dead of disease (DOD) | 10 | 10.10 |  |  |  |
| **Infiltration** | N | % | **Lymphatic Vessels Invasion** | N | % |  |  |  |
| Espansive | 49 | 49.49 | Absent/focal | 72 | 72.73 |  |  |  |
| Infiltrative | 50 | 50.51 | Substantial | 27 | 27.27 |  |  |  |
| **Desmoplasia** | N | % | **Blood Vessels Spaces Invasion** | N | % |  |  |  |
| Absent | 20 | 20.20 | Absent | 89 | 89.90 |  |  |  |
| Present | 79 | 79.80 | Present | 10 | 10.10 |  |  |  |
| **Intratumoral Necrosis** | N | % | **Tumor Infiltrating Lymphocytes (TIL)** | N | % |  |  |  |
| Absent | 37 | 37.37 | Absent | 74 | 74.75 |  |  |  |
| Present | 62 | 62.63 | Present | 25 | 25.25 |  |  |  |

The mean age of the patient was 68,49±11,26 and they were mainly affected by EEC (77.8% of the total cases) and by early stage disease (FIGO I-II) (73.7% of the total cases). We found 69 low-grade EEC, 8 high-grade EEC and 22 NEEC.

For the IHC staining indexes, in our study population we observed a fair strong and significant correlation between ER and PR (rho=0.668; p<0.001) and between e-cadherin and β-catenin (rho=0.671; p<0.001); a weaker but significant correlation was observed between ki67 and p53 (rho=0.531; p<0.001). We observed a significant inverse correlation between PR and Ki67 (rho=-0.457; p<0.001) and between PR and p53 (rho=-0.436; p<0.001) (Additional Material 2).

### 2-Univariate Analysis

Factors that proved significantly correlation with a worse disease free survival (DFS) were: NEEC, substantial LVSI, FIGO III-IV. An infiltrative pattern of invasion and presence of TIL showed a borderline significance (p=0.06 and p=0.008 respectively). No patients with absent intratumoral necrosis had recurrent disease (Table 3).

**Table 3.** Univariate Analysis: histopathological prognostic factors.

|  |  | OS |  |  | DFS |  |  |
| --- | --- | --- | --- | --- | --- | --- | --- |
|  |  | p | HR | 95% C.I. | p | HR | 95% C.I. |
| Histology | EEC<br>NEEC | **0.0005** | 13.7169 | 3.1554-59.6286 | **0.0002** | 10.4586 | 3.0125-36.3096 |
| Stage (FIGO) | FIGO I-II<br>FIGO III-IV | 0.0861 | 3.2927 | 0.8445-12.8385 | **0.0071** | 4.6933 | 1.5216-14.4763 |
| Infiltration | Expansive<br>Infiltrative | 0.0806 | 3.0901 | 0.8716-10.9549 | 0.0646 | 2.615 | 0.9435-7.2475 |
| Desmoplasia | Absent<br>Present | **0.0319** | 5.1795 | 1.1530-23.2676 | 0.2425 | 2.1039 | 0.6044-7.3242 |
| Intratumoral<br>necrosis | Absent<br>Present | 0.0246 | NA | NA | 0.0025 | NA | NA |
| TIL | Absent<br>Present | 0.1558 | 3.0336 | 0.6552-14.0452 | 0.07809 | 1.1854 | 0.3576-3.9300 |
| LVSI | Absent<br>Present | **0.0507** | 3.8517 | 0.9958-14.8985 | **0.0069** | 4.7367 | 1.5335-14.6305 |

Regarding IHC, cases with low PR and high Ki67 demonstrated an increased recurrent disease HR, statistically significant. ER-correlation reached only a borderline significant value (p=0.08) (Table 4).

**Table 4.** Univariate Analysis: immunohistochemical prognostic factors.

|  |  | OS |  |  | DFS |  |  |
| --- | --- | --- | --- | --- | --- | --- | --- |
|  |  | p | HR | 95% C.I. | p | HR | 95% C.I. |
| ER | High<br>Low | 0.0648 | 4.6526 | 0.9097-23.7951 | 0.0811 | 3.3735 | 0.8605-13.2255 |
| PR | High<br>Low | **0.0174** | 5.5003 | 1.3487-22.4316 | **0.0140** | 4.3018 | 1.3433-13.7767 |
| p53 | Wild-type<br>Abnormal | 0.0804 | 3.3348 | 0.8645-12.8646 | 0.2056 | 1.9716 | 0.6891-5.6410 |
| Ki67 | High<br>Low | **0.0231** | 4.27 | 1.2210-14.9931 | **0.0340** | 3.0195 | 1.0871 to<br>8.3869 |
| β-catenin | High<br>Low | 0.2237 | NA | NA | 0.1440 | NA | NA |
| e-cadherin | High<br>Low | 0.2840 | NA | NA | 0.7799 | 1.2937 | 0.2125-7.8751 |
| cyclin-D1 | High<br>Low | 0.9737 | 1.0213 | 0.2874-3.6291 | 0.8341 | 1.119 | 0.3908-3.2039 |
| bcl-2 | High<br>Low | 0.6108 | 1.0217 | 0.2845-3.6686 | 0.9341 | 1.045 | 0.3680-2.9673 |

Concerning overall survival (OS), NEEC, presence of desmoplasia and LVSI were correlated with an unfavourable OS, while FIGO III-IV and infiltrative pattern of invasion only with borderline significance (p=0.09 and p=0.08 respectively); in analogy to DFS no patients without intratumoral necrosis died of the disease (Table 3).

Regarding IHC, cases with low PR, high Ki67 demonstrated an increased HR of DOD, while ER and p53-correlation reached only a borderline significant value (p=0.06 and p=0.08 respectively) (Table 4).

### 3-Multivariate Overall Survival Analysis

In our study population, the multivariate DFS analysis based on the Cox-Model evidenced a fair strong and significant inversely correlation between the survival function and tumor stage (p=0.0042). Even if an inversely relation between poor DFS and NEEC, presence of necrosis and high ki67 expression covariates was observed, it did not reach a statistical significance (Table 5).

**Table 5.** Multivariate Analysis (OS and DFS): analysis of parameter estimates.

| OS |  |  |  |  |  |  |
| --- | --- | --- | --- | --- | --- | --- |
|  | DF | Estimate | Standard Error | Chi-Square | Pr > ChiSq | Hazard Ratio |
| Stage | 1 | -1.114 | 0.3097 | 12.934 | 0.0003 | 0.3283 |
| Histology | 1 | -0.011 | 0.3206 | 0.0012 | 0.9725 | 0.989 |
| Desmoplasia | 1 | -0.6529 | 0.3144 | 4.3132 | 0.0378 | 0.5205 |
| Necrosis | 1 | 0.0513 | 0.2715 | 0.0357 | 0.8501 | 1.0527 |
| PR | 1 | 0.4458 | 0.3181 | 1.9645 | 0.161 | 1.5618 |
| Ki 67 | 1 | -0.5401 | 0.2781 | 3.7715 | 0.0521 | 0.5827 |
|  | -2 Log Likelyhood | Chi-Square | DF | Pr > ChiSq |  |  |
|  | 600.8455 | 21.4348 | 6 | 0.0015 |  |  |

| DFS |  |  |  |  |  |  |
| --- | --- | --- | --- | --- | --- | --- |
|  | DF | Estimate | Standard Error | Chi-Square | Pr > ChiSq | Hazard Ratio |
| Stage | 1 | -1.0175 | 0.3555 | 8.1926 | 0.0042 | 0.3615 |
| Histology | 1 | -0.0434 | 0.3526 | 0.0151 | 0.9021 | 0.9575 |
| Necrosis | 1 | -0.0525 | 0.2728 | 0.037 | 0.8474 | 0.9488 |
| LVSI | 1 | 0.026 | 0.3342 | 0.006 | 0.938 | 1.0263 |
| PR | 1 | 0.2247 | 0.3288 | 0.4672 | 0.4943 | 1.252 |
| Ki 67 | 1 | -0.3806 | 0.2844 | 1.7915 | 0.1807 | 0.6834 |
|  | -2 Log Likelyhood | Chi-Square | DF | Pr > ChiSq |  |  |
|  | 558.4809 | 16.3529 | 6 | 0.012 |  |  |

The multivariate OS analysis evidenced a fair strong and significant inversely correlation between survival and tumor stage (p<0.001) and presence of desmoplasia (p=0.05). Ki67 expression showed a significant borderline correlation (p=0.0521) with the survival function. The hemi-life survival time (2^nd^ quartile) was 28 months (Table 5).

## Discussion

We proposed a selected number of histopathological and IHC markers, relatively inexpensive and reproducible even in peripheral centers.

Although the limited case study, low-grade EEC showed to be the most prevalent histotype according to previous literature.^2^

As stated above, the role of many molecular markers has been investigated in the last years and TGCA identified four categories with a good correlation with prognosis, potentially superior to histopathological features. However the last studies demonstrated that this molecular classification does not replace clinical and pathological risk assessment, thus requesting a strict integration.^7^

Some of the proposed histological and IHC prognostic factors confirmed their role in term of OS and DFS survival.

Particularly, the advanced stage at diagnosis appeared to be the most crucial prognostic element. Indeed stage is the only parameter – we investigated – which maintains a strong significance correlation with OS and DFS in multivariate analysis. Moreover, previous studies showed stage significant prognostic value, also independently from molecular classification.^20^

In our cases, NEEC mostly consisted of serous carcinoma, even intermingled with other histotypes to determine mixed substypes. These patients showed a higher recurrence rate in univariate analysis, confirming a worse prognosis as well as described in previous works.^28^

Regarding morphological parameters, LVSI - described as presence of neoplastic cells within endothelial lined space - is recently classified as absent/focal and substantial/extensive (≥3 lymphvascular spaces).^7,29^As stated in literature, LVSI has a great prognostic impact when substantial (extensive), associate with a significant increase in the risk of recurrence.^30,31^ In our study substantial LVSI showed a significant worse prognostic value for recurrence and overall survival. Even if the LVSI lost its statistical significance in multivariate analysis, it still remains an essential risk parameter, mandatory to be underlined in pathological report.

In our study desmoplasia resulted significantly correlated with a poor prognosis (only OS) after multivariate analysis. However univocal dates about desmoplasia prognostic role are not available^32^ and the criteria defining desmoplasia are still not clearly established; particularly we evaluated the fibroblastic reaction (hematoxylin-eosin stain) at the tumor edge.

Tumor necrosis has been associated with aggressive features and poor prognosis.^33^ We did not observe any relapsing or deadly disease in patients affected by EC with no intratumoral necrosis.

Among morphological features, TIL is usually associated with favorable outcome both in EEC and NEEC.^34^ However in our cases TIL did not achieve a full significant prognostic correlation. This data may be affected by the little cohort and the method of lymphocytes count only on HE stain without any IHC marker, with subsequent possible underscoring.

Regarding IHC parameters, many different studies investigated the prognostic role of hormonal receptors status in the last 30 years. A meta-analysis confirmed the better prognosis of ER+ and PR+ EC.^35^ However, despite the large data on this topic, we observed heterogeneity in techniques and evaluation of ER and PR expression. If low levels of PR have been invariably associated to higher grade, stage, ki67, p53 and non-endometrioid histology and, as consequence to a worse DFS and OS,^12,36^ the clinical significance of ER is much more debated.^10^

In our case study we observed low PR expression as poor prognostic factor (univariate analysis), whereas only a borderline correlation between low ER expression and poor prognosis. However we evidenced a direct trend between ER and PR, being the latter low expression related to high ki67 index and p53 overexpression (see Additional Material 2), remarking its unfavorable value.

Mutation of TP53 are par excellence related to aggressive tumors. It is a crucial and early event in the development of serous carcinoma. However, its expression may also appear in high grade EEC and in a small percentage of low grade EEC, being alone not sufficient for a proper differential diagnosis of serous subtype.^37^ p53 IHC marker showed to be a good surrogate of the molecular TP53 mutation with a high sensitivity and specificity.^38^ Several studies have demonstrated a significant correlation between TP53 mutation and a worse clinical outcome, independent of histotype and grade.^31,39^ We scored the percentage of positive cells as wild type expression and aberrant/mutation type.

Despite the correlation with aggressive tumor forms, our study found that mutations in TP53 are not significantly related to a reduction in OS and DFS.

A high proliferation index (%Ki67) significantly and inversely correlates with OS in multivariate analysis, but the same parameter does not reach statistical value when correlated with DFS.

Studies report cyclin D1 expression as a prognostic factor, since its overexpression has been correlated with the neoplastic lymph node involvement.^29^ On the contrary, in our study it does not appear related to prognosis (both OS and DFS). Also bcl-2 seems to be an independent prognostic factor. Some studies show the close correlation between its loss and lymph node metastasis and relapsing disease.^23,24,25^ We did not find a significant influence of this marker on OS and DFS.

β-catenin shows a membranous expression in normal endometrium, while CTNNB1 mutation drives to a β-catenin membranous staining reduction together with its nuclear accumulation. In one study,^40^ β-catenin expression resulted associated with endometrial hyperplasia-carcinoma sequence and β-catenin membranous staining loss was evidenced alongside the transition from hyperplasia to high grade EEC. Another study identified nuclear β-catenin expression as poor prognostic factor (DFS) when only considering the subset of low-grade EEC. However β-catenin progressive nuclear accumulation seems to be variedly reliable when used as CTNNB1 mutation surrogate.

In our study, membranous β-catenin expression did not result determinant for OS and DFS, in part probably due to tumor grading non-selection or due to merely membranous IHC evaluation.

Likewise, loss of e-cadherin has also been considered as independent prognostic factors, related to a more aggressive behavior.^22,26^ In our case e-cadherin did not demonstrate to be prognostically determinant, even if we found a positive and direct trend between its expression and membrane staining of β-catenin, both confirming their primary function as adhesion molecules. (see Additional Material 2).

## Conclusion

The pathological report still remain an essential step for patient’s risk stratification. Morphological features as histological subtype, LVSI and stage evaluation should be necessarily specified. Desmoplasia, even understated and not univocally defined, may be worth to be reported for its prognostic value. However further investigations and a larger number of cases are needed to frame this issue.

Our limited study population could have affected the prognostic evaluation of some IHC markers, generally considered as significant related to outcome. However, among relevant IHC markers, hormonal receptors and ki67 could embed clinical information in order to stratify patients’ risk, even regardless of the recent TGCA classification, not always easily available in routinely lab sets.

Joint efforts are focusing on the development of sensitive and specific molecular IHC surrogates, which provide the best patient tailored care and targeted therapy.

## Data Availability

All the data concerning the presented study, in anonimized form are available to the asking scholars

## Abbreviations

EC: endometrial carcinoma
EEC: endometrioid endometrial carcinoma
NEEC: non endometrioid endometrial carcinoma
TCGA: Cancer Genome Atlas Research Network
IHC: immunohistochemistry
ER: estrogen receptor
PR: progesterone receptor
HE: hematoxylin-eosin stain
SI: staining index
DOD: dead of disease
DFS: disease free survival
HR: hazard ratio
OS: overall survival
LVSI: lymphovascular spaces invasion
TIL: tumor infiltrating lymphocytes

## Acknowlegements

The authors have declared no conflicts of interest.

The current study was founded with University of Genoa grants (FRA).

Special thanks to Giorgia Anselmi, Silvana Bevere, Simona Pigozzi and Laura Zito for technical assistance.

## Author contributions

Michele Paudice and Giulia Scaglione, wrote paper; Chiara Maria Biatta, performed research study; Marianna Riva, Bruno Spina and Gabriele Gaggero collecting data; Ezio Fulcheri, analyzed data; Simone Ferrero and Fabio Barra, gynecological correlations; Valerio Gaetano Vellone, designed study and wrote paper.

