## Additional Tables 1 & 2 for "Multivariate Analysis Of Histopathological And Immunohistochemical Prognostic Factors In Endometrial Carcinoma. A Retrospective Pilot Study Of An Italian Regional Referral Center"

Additional Material 1

IHC markers values distribution. The percentage of positive cancer cells is expressed on the X-axis while the relative frequency in the study population is expressed in Y-axis. The red line graphically expresses the dichotomy in the study population.

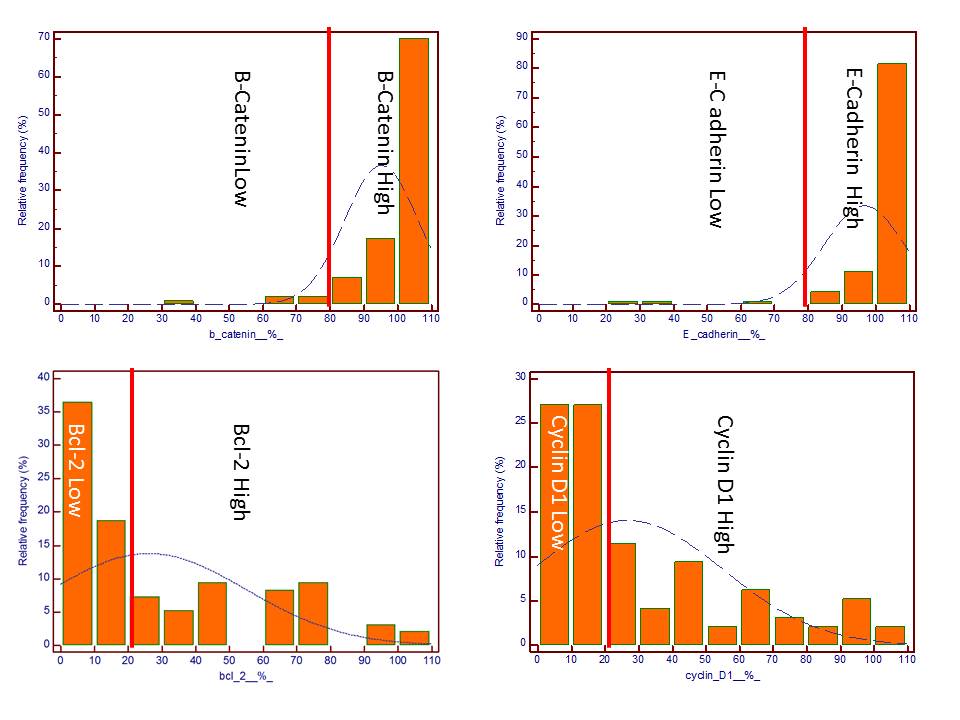

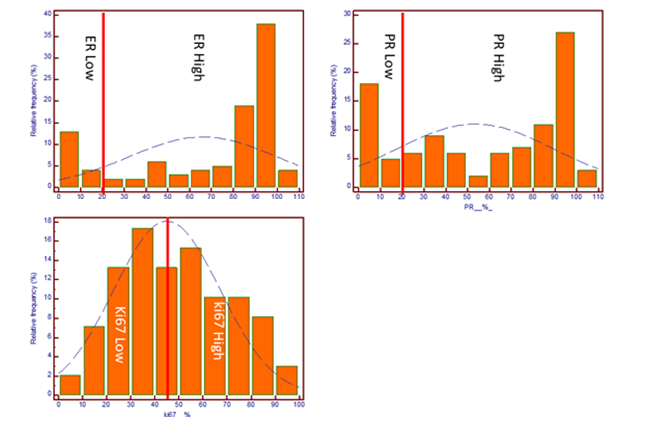

Additional material 2: Correlation between continuous variables.

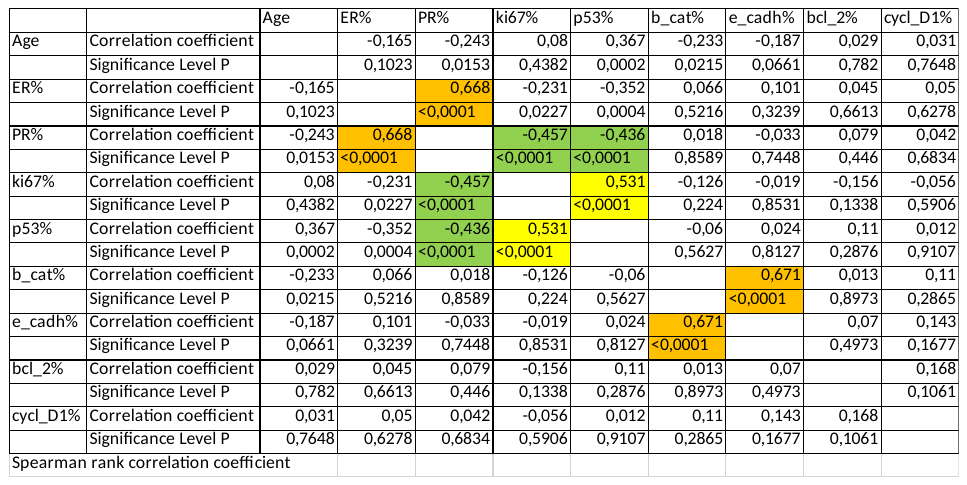
